# Developing and Prospectively Validating a Reproducible Graph Representation Specification for Clinical Guideline Algorithms: The Measurement Foundation of the Clinical Guideline Complexity Index

**DOI:** 10.64898/2026.07.17.26358358

**Authors:** Richard V. Milani, Robert M. Bober

## Abstract

**Background:** Translating a clinical guideline decision algorithm into a computational graph requires judgment, and unconstrained coding yields divergent graphs; any complexity measure computed from such a graph inherits that variation, so its reproducibility must be demonstrated rather than assumed.

**Objective:** To develop, and prospectively test, an empirical method for making graph extraction reproducible, using the Clinical Guideline Complexity Index (CGCI) and four guideline algorithms as a case study.

**Methods:** We built a Graph Representation Specification (an ontology, a motif catalogue, disambiguation conventions, decomposition rules, a deterministic validator, and a scoring engine) and refined it by error-driven grammar induction: measure inter-coder disagreement, localize its dominant class, induce a single grammar rule, and prospectively test whether that rule improves agreement in the anticipated class. Reproducibility was quantified with a pre-specified, topology-based endpoint (Decision Topology Agreement) rather than edge agreement, which is oversensitive to representational choices that do not affect the score. Two trained coders independently coded the diabetes, dyslipidemia, heart-failure, and hypertension algorithms.

**Results:** A rule induced from the diabetes comorbidity panel (assessment topology) generated a pre-specified prediction that heart-failure figures, sharing the same motif, would converge; on a fresh, independently coded pair they did, with an absolute CGCI difference of approximately one. Decision topology reproduced closely (decision-order agreement at or near 1.00 for three of four guidelines), while breadth counting was rule-sensitive: an explicit modifier-counting rule reduced the largest disagreement from 27 to 4 tokens. Residual disagreement was bounded and localizable to specific, nameable representational choices.

**Conclusions:** Graph-extraction reproducibility can be systematically improved through iterative grammar refinement, and a prospectively derived rule can be confirmed to improve agreement. These results establish the measurement foundation (reliability, not construct validity) for a companion study interpreting CGCI as cognitive load, and the method may apply wherever graphs are extracted from structured source artifacts.

**Highlights:**

- A reproducible method converts guideline decision figures into scorable graphs.
- Coder disagreement is localized, encoded as a rule, and prospectively tested.
- A rule induced from diabetes prospectively improved heart-failure agreement.
- Decision topology reproduced closely; breadth counting needed explicit rules.
- A frozen specification lets an independent team implement the parser.

## 1. Introduction

Whenever a person translates a structured visual artifact into a computational representation, judgment enters. Graphs are increasingly the computational substrate for clinical decision support, workflow automation, and automated reasoning, so the reproducibility of graph extraction is a practical concern and not only a theoretical one [1,2]. A flowchart, a decision tree, or a clinical algorithm does not map onto a graph in one canonical way: readers differ in where they place a decision, how finely they decompose a step, and which annotations they treat as inputs rather than outcomes. If a representation is to support quantitative analysis, this judgment must be constrained without being eliminated: the representation should remain interpretable yet be produced reproducibly by different coders. Reproducible graph grammars offer one route to that goal, and how to develop such a grammar is the problem this paper addresses.

Clinical guideline decision algorithms are a demanding instance of the problem. Modern guidelines increasingly present management as branching figures (comorbidity-driven pathways, tiered drug selections, threshold ladders, and reassessment loops) that vary widely in structural complexity. Quantifying that complexity reproducibly is a prerequisite for a range of downstream uses, including estimating clinician workload, studying cognitive load, and designing decision support [3]. A measure of algorithmic complexity is only as trustworthy as the representation it is computed from; if two trained readers encode the same figure into materially different graphs, any complexity score inherits that variation.

The Clinical Guideline Complexity Index (CGCI) is one such measure, computed from a graph representation of a guideline algorithm and combining a multiplicative branching-depth core with additive terms for decision variables and modifiers. Unlike prose-oriented readability or complexity instruments [9–11], CGCI is a structural measure: it depends on the topology of the decision graph rather than on the language of the guideline. This makes reproducibility a question about the graph, not the text, and therefore a question that must be demonstrated rather than assumed. Computer-interpretable guideline formalisms have been developed to encode guidelines for execution [4–8], but these focus on making a guideline executable rather than on the reproducibility of the human coding that produces the representation.

A natural way to assess graph reproducibility is to compare the edges two coders draw. We found this endpoint to be misleading, because minor representational choices (how a shared downstream pathway is wired, or whether a reassessment loop is drawn explicitly) change individual edges without changing the decision topology that determines the score. Edge agreement thus penalizes irrelevant variation. We therefore pre-specified a topology-based endpoint, Decision Topology Agreement, that compares codings at the level of decisions and their ordering rather than raw edges.

The central contribution of this work is not a new complexity metric; it is a generalizable methodology for converting structured visual decision algorithms into reproducible graph representations suitable for quantitative analysis. Concretely, it is a Graph Representation Specification (an ontology, a motif catalogue, disambiguation conventions, decomposition rules, a validator, and a scoring engine) together with an empirical method for developing it that we term error-driven grammar induction: measure inter-coder disagreement, localize its dominant class, induce a single grammar rule that resolves it, and prospectively test whether that rule improves agreement in the anticipated class. CGCI and four clinical guidelines are the motivating case study through which the specification and method are developed and evaluated; the method itself is not specific to guidelines and may apply to other settings in which graphs are extracted from structured source artifacts.

We pose three pre-specified questions:

**RQ1.** Can graph-extraction reproducibility be systematically improved through iterative grammar refinement?
**RQ2.** Does a prospectively derived grammar rule improve agreement in the anticipated class of disagreement?
**RQ3.** After refinement, which aspects of the graph representation remain the principal sources of disagreement?

Finally, this paper occupies a defined position relative to a companion study. As summarized in Figure 1, a guideline passes through the specification to a validated graph and a CGCI score, which the companion paper then interprets in terms of cognitive load. The present paper concerns everything up to and including the score: whether the representation and the score are reproducible. It establishes reliability; it does not, and does not claim to, establish that the score measures cognitive load. That construct-validity question is the companion paper’s subject, and the two papers together form a reliability-plus-validity argument. The companion study applies the CGCI framework across a broader set of guideline iterations spanning a decade and interprets the resulting scores in terms of cognitive load; its analysis rests directly on the score reproducibility established here.

**Figure 1.**
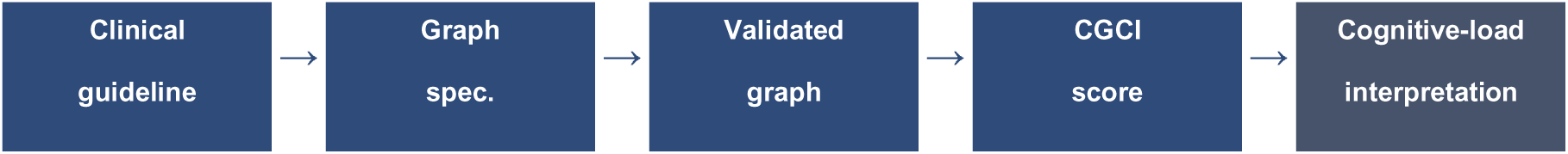
A guideline algorithm is coded through the Graph Representation Specification into a validated graph and a CGCI score; a companion paper interprets that score in terms of cognitive load. This paper covers the first four stages and not the final interpretive step.

The refinement method at the heart of this paper is summarized in Figure 2 and specified in the Methods; we introduce it here because the reproducibility results, the residual analysis, and the future-work plan all follow from it.

**Figure 2.**
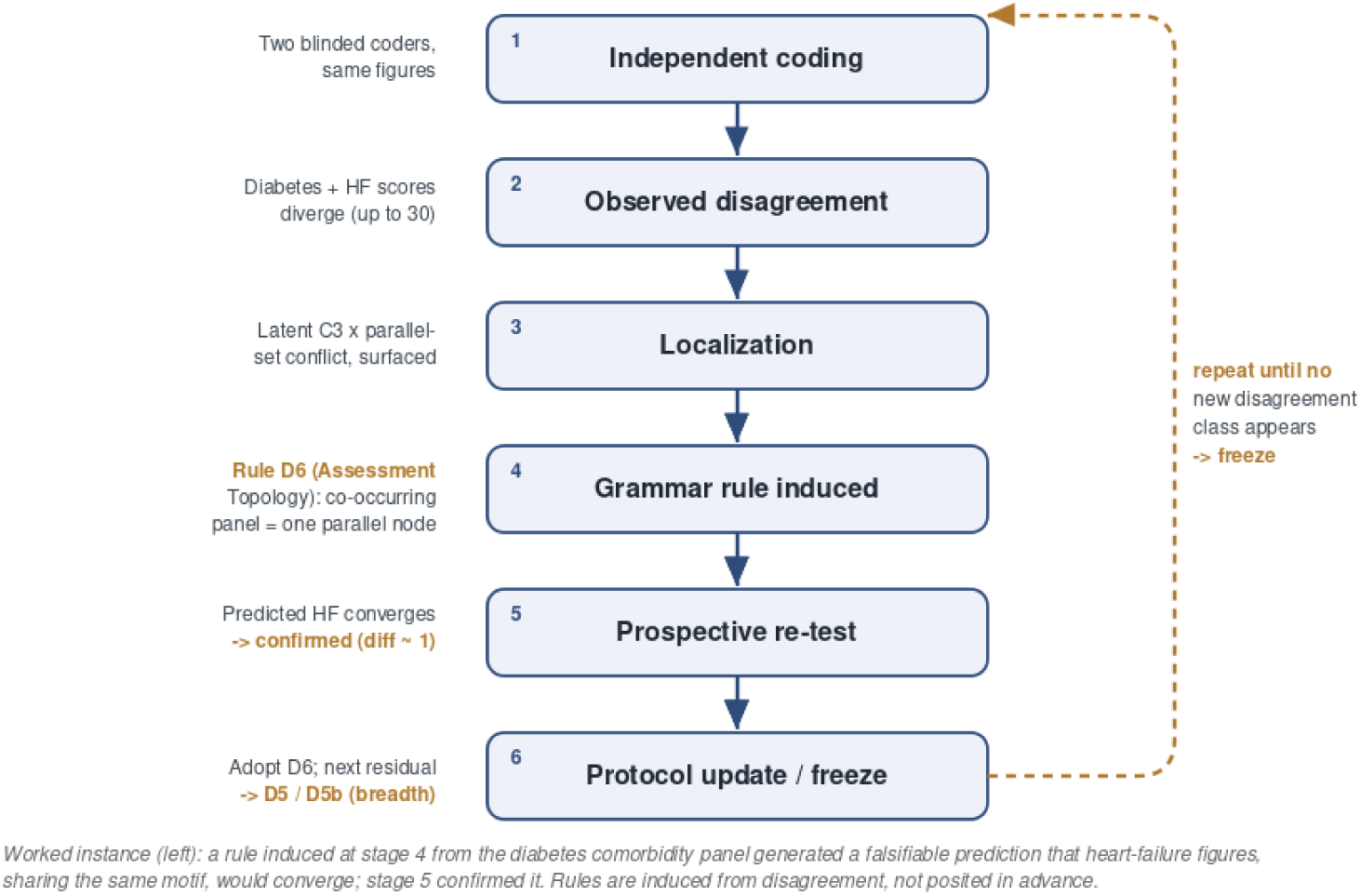
Error-driven grammar induction. Each turn induces one rule from observed disagreement and prospectively tests it; the worked instance at left is the D6 cycle.

## 2. Methods

### 2.1 The complexity measure

The Clinical Guideline Complexity Index (CGCI) summarizes the structural complexity of a guideline decision algorithm from a graph representation of that algorithm. For a given algorithm, CGCI = (B × D) + V + M, where B is the number of branch points (decision nodes with two or more distinct successors), D is decision depth (the mean number of decision nodes encountered along root-to-leaf paths, computed after removing feedback edges), V is the number of distinct decision variables, and M is the number of distinct modifiers. Two further quantities (the number of terminal pathways and a topological-phase indicator) are recorded and reported alongside the composite rather than summed into it. The multiplicative B × D core couples breadth and depth so that an algorithm that is both wide and deep is scored super-additively, reflecting the compounding demand of nested branching, while V and M contribute additively as breadth-of-input terms [12]. This structure is a deliberate design choice rather than an empirically fitted model.

### 2.2 The Graph Representation Specification

Translating a published guideline figure into a scorable graph is judgment-laden, and unconstrained coding yields divergent graphs even among trained readers. We therefore developed a Graph Representation Specification in which a formal grammar is one component among several; throughout, we use “specification” for the whole and reserve “grammar” for the rule layer within it (which the ontology, conventions, validator, and scoring engine surround). The specification comprises: an ontology defining three node types (decision, action, and terminal) together with directed edges and node attributes (variables on decisions; modifiers on decisions, actions, or terminals; and a terminal class on terminals); a catalogue of five recurring structural motifs (the sequential gate, parallel assessment, shared rail, threshold ladder, and reassessment loop), each with an explicit recognition test and a single canonical encoding; a set of disambiguation conventions; a set of decomposition rules governing granularity; a deterministic validator that rejects structurally invalid graphs; and a scoring engine that computes CGCI from a valid graph. A motif is a recurring local graph topology that admits a single canonical representation. Coders first classify the local graph motif of a figure region before drawing nodes, because the motif fixes the local topology and the topology governs the finer decomposition choices. The complete normative specification, sufficient for an independent team to implement the parser, is given in Appendix A.

Three decomposition rules, each a product of the refinement process described below, are central to reproducibility. Rule D6 (assessment topology) specifies that a set of indicators that can co-occur in one patient and are each evaluated independently is encoded as a single assessment decision carrying each indicator as a variable and branching in parallel, rather than as a chain of sequential gates [8]. This choice has a quantitative rationale: encoded as a sequential cascade (with the implicit negative edges added by Convention C3), a panel of co-occurring indicators feeds the multiplicative B × D core, so that in this representation its contribution grows approximately quadratically with the number of indicators; encoded as a single parallel-assessment decision under D6, the same panel feeds the additive variable term and grows approximately linearly. Section 3.1 and Figure 4 show this inflation empirically. Rule D5 (semantic decomposition) specifies that variables are counted at the granularity the figure itself labels: a composite term or a validated score is a single variable and is not expanded into implicit constituents. Rule D5b (modifier counting) distinguishes a modifier, which qualifies a recommendation (a dose caveat, intensity or efficacy tier, numeric target, contraindication, monitoring cadence, or population qualifier), from a distinct clinical action such as a risk calculation, screening step, referral, or procedure, which is encoded as its own node rather than as a modifier.

### 2.3 Error-driven grammar induction

The specification was not specified a priori; its rules were induced from observed disagreement through an iterative cycle (Figure 2). In each iteration, two coders independently coded the guideline set; inter-coder disagreement was measured and localized to a dominant class; a single grammar rule addressing that class was added; the revised rule generated a prospective, falsifiable prediction about which figures should subsequently converge; and a fresh independent coding was used to test that prediction. The cycle terminates (the protocol is frozen) when successive coding cohorts no longer surface a new disagreement class requiring a new rule. This error-driven induction of grammar rules is analogous to refinement procedures in natural-language processing, compiler construction, and knowledge engineering, in which production rules are derived from observed failures rather than posited in advance [13–16]. Unlike those fields, however, the production rules here are induced from observed human disagreement rather than optimized for machine parsing.

### 2.4 Reproducibility endpoint

Representation-level agreement was quantified with a pre-specified metric, Decision Topology Agreement (DTA). We did not adopt edge-level agreement as the primary endpoint: minor representational choices (how a shared rail is wired, or whether a reassessment loop is drawn explicitly) alter individual edges without changing the decision topology that determines CGCI, so edge agreement penalizes representational variation that is irrelevant to the score. DTA instead compares two codings after contracting each graph to its decision nodes, and reports three components: agreement in the number of decisions, the rate at which decisions are matched between coders (by an anchor-first, otherwise label-similarity, optimal assignment), and agreement in the relative order of matched decisions. The metric is deterministic and invariant to which coder is designated first. Agreement on the breadth attributes (variables and modifiers) is reported separately, because the structural and breadth layers of the representation behave differently.

### 2.5 Study design and analysis

Four guideline decision algorithms were coded: the ADA diabetes glucose-lowering and individualized-target algorithms [17], the ACC/AHA cholesterol algorithms [18], the AHA/ACC/HFSA heart-failure algorithms [19], and the ACC/AHA hypertension algorithms [20]. Two trained coders coded independently and blind to one another using a figure-anchored data-entry workbook, in which each decision is tagged with the figure’s own printed step or box label to support cross-coder alignment. Graphs were scored with a deterministic engine and compared with the DTA procedure. Reproducibility is assessed as agreement between independent coders rather than as accuracy against a gold standard; no consensus reference graph is used, and the canonical encoding for any figure region is defined by the specification (Appendix A). Because the design comprises four algorithms and two coders, we report per-guideline agreement and component-level differences rather than a single pooled reliability coefficient, which would carry an uninformatively wide confidence interval at this sample size [21]; the study is framed as measurement development rather than psychometric validation [22,23], and a confirmatory study using the frozen protocol and an expanded guideline set is pre-registered as future work.

## 3. Results

### 3.1 Prospective validation of protocol refinement

The central result is that a grammar rule induced from one guideline’s disagreement prospectively improved agreement on a different guideline, in the class of figure the rule was designed to address. Across three protocol versions, the mean absolute CGCI difference on the two guidelines under active development (diabetes and heart failure) moved from 12.8 to 30.2 to 6.7 (Table 1). The increase at the intermediate version was diagnostic: a convention that added an implicit negative edge to eligibility decisions, when applied to a panel of co-occurring comorbidities, chained them into a sequential cascade that inflated both branch count and depth (Figure 4). Localizing this failure produced Rule D6 (assessment topology), which encodes such a panel as a single parallel-assessment decision. D6 generated a pre-specified prediction: heart failure, whose device- and therapy-selection panels share the same parallel-assessment motif as the diabetes comorbidity panel, should converge under the revised rule. On a fresh, independently coded pair, the heart-failure absolute CGCI difference fell to approximately 1.0, and the diabetes topology converged with the residual isolated to variable and modifier granularity, confirming the prediction and answering RQ2. This sequence constitutes prospective evidence that disagreement-driven rule induction can improve graph reproducibility.

**Table 1.** Mean absolute CGCI difference between coders on the two guidelines under active development, across three protocol versions. The rise at v2.4 is diagnostic; localizing it produced Rule D6, whose prospective prediction is tested in the text.

| Protocol version | Change introduced | Diabetes + HF mean<br>\|ΔCGCI\| |
| --- | --- | --- |
| v2.3 | baseline | 12.8 |
| v2.4 | added convention C3 | 30.2 |
| v2.5 | added Rule D6 | 6.7 |

### 3.2 Decision topology is reproducible

Under the frozen protocol, with figures coded using anchor tags, decision topology reproduced closely between coders (Table 2, Figure 3A). Decision-order agreement (whether two coders that matched a decision agreed on its position in the algorithm) was at or near 1.00 for three of the four guidelines, and 0.70 for the fourth; decision-node match rates ranged from 0.67 to 1.00 and were highest where anchor tags aligned the coders’ labels, reaching 0.95 for the most complex guideline (dyslipidemia, with 21 to 22 decisions). For hypertension the two codings were topologically identical. These results suggest that the structural core of CGCI, the B × D term, is more reproducible than the breadth components under the current protocol; it is also the component most relevant to any downstream comparison of complexity across guidelines.

**Figure 3.**
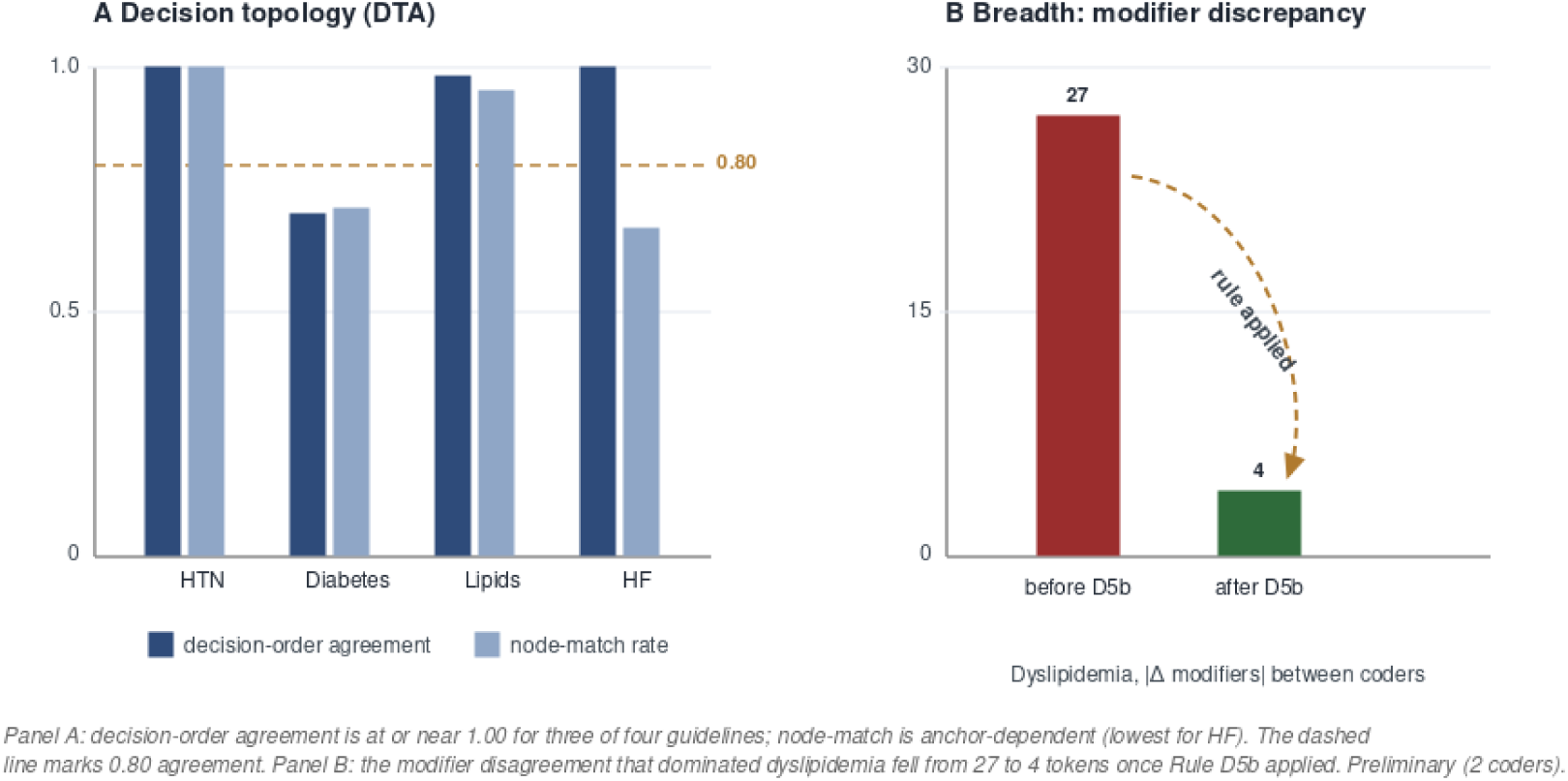
Two-level reproducibility. (A) Decision-topology agreement per guideline. (B) The dyslipidemia modifier disagreement before and after Rule D5b. Preliminary (two coders).

**Figure 4.**
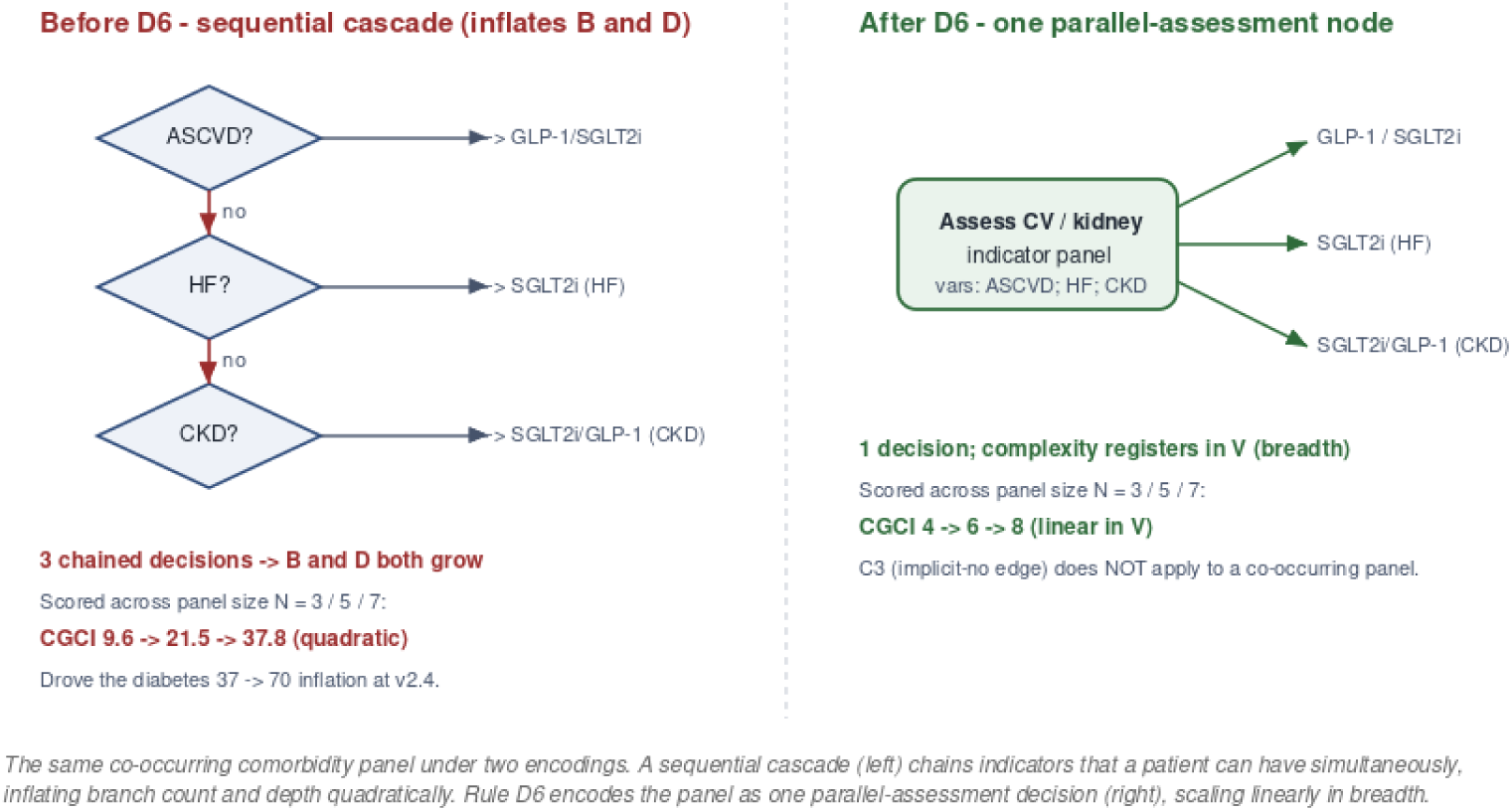
Two encodings of the diabetes comorbidity panel. A sequential cascade (left) inflates branch count and depth quadratically; Rule D6 encodes the panel as one parallel-assessment decision (right), scaling linearly in breadth.

**Table 2.** Decision Topology Agreement components between the two coders, per guideline, under the frozen protocol with figure anchoring. Figures are preliminary (two coders, four guidelines).

| Guideline | Decisions (A / B) | Count agr. | Node-match | Order agr. |
| --- | --- | --- | --- | --- |
| Hypertension | 7 / 7 | 1.00 | 1.00 | 1.00 |
| Diabetes | 8 / 6 | 0.75 | 0.71 | 0.70 |
| Dyslipidemia | 21 / 22 | 0.96 | 0.95 | 0.98 |
| Heart failure | 5 / 4 | 0.80 | 0.67 | 1.00 |

### 3.3 Breadth counting is rule-sensitive, and the rules resolve it

Agreement on breadth attributes was initially poorer than on topology and was the dominant remaining source of score disagreement once topology had converged. The largest discrepancy, on dyslipidemia, arose because one coder recorded separate clinical steps (risk calculation, cascade screening, genetic testing, specialist referral) as modifiers while the other recorded only therapy qualifiers. Rule D5b (modifier counting) draws this distinction explicitly. After its introduction, the dyslipidemia modifier discrepancy fell from 27 to 4 tokens and the corresponding CGCI difference to 3.9; the diabetes difference fell to 1.9 (Table 3, Figure 3B). This demonstrates that breadth disagreement is not irreducible noise but is resolved by an explicit counting rule of the same kind produced elsewhere by the refinement cycle.

**Table 3.**
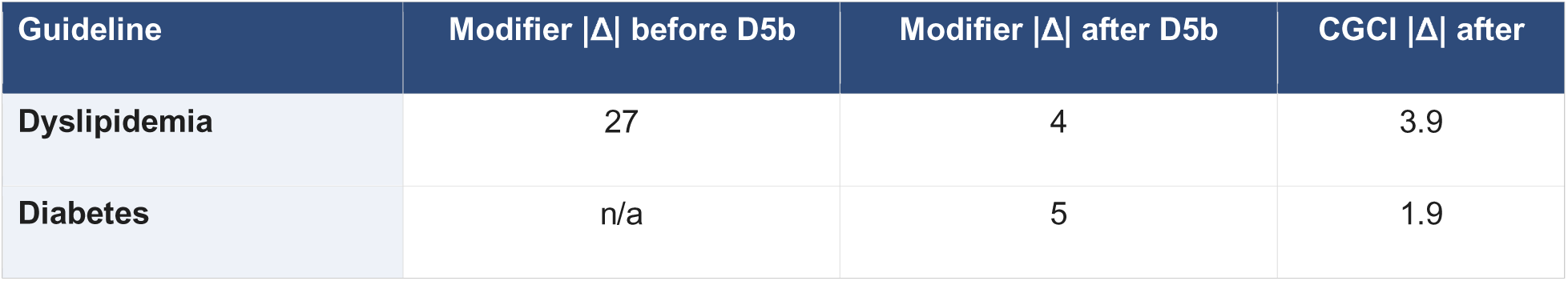
Effect of Rule D5b (modifier counting) on the two guidelines where the modifier layer dominated disagreement.

### 3.4 Residual disagreement is bounded and localizable

Under the frozen protocol the mean absolute CGCI difference across the four guidelines was 7.6. The residual did not diffuse across the representation but localized to two nameable sources: incomplete modifier recording on hypertension (one coder omitted dosing modifiers that the other captured) and the coding of eligibility thresholds (for example, the ejection-fraction and QRS thresholds that gate device therapy), which one coder recorded as modifiers and the other as decision inputs. The latter was resolved by specifying that eligibility thresholds are variables of the candidacy decision. That every residual could be named and assigned to a specific representational choice, rather than attributed to general coder variability, is itself evidence that the specification localizes disagreement, the property that drives the refinement cycle.

### 3.5 Structure and breadth vary independently

Hypertension illustrates the separability of the specification’s layers. The two codings produced identical B × D (7 × 1.6) and identical decision topology (DTA of 1.00), yet differed in CGCI; the entire difference arose from the modifier count. Structural complexity and breadth of inputs are therefore measured independently, and a disagreement in one does not imply a disagreement in the other. This separation is what allows the reproducible structural core to be reported with confidence even where the breadth layer still varies. More broadly, localization of residual disagreement, rather than its elimination, is the defining property of the specification.

## 4. Discussion

Across four clinical guideline algorithms and two independent coders, decision topology reproduced closely, while breadth attributes required explicit counting rules to converge. More importantly, the process by which those rules were obtained was itself testable: a rule induced from the diabetes comorbidity panel generated a prospective prediction that heart-failure figures would converge, and that prediction held. The paper’s primary finding is therefore methodological (that graph-extraction reproducibility can be improved by error-driven grammar induction, and that a rule so induced can be prospectively confirmed), with the clinical measure serving as the vehicle for the demonstration.

### 4.1 Why the process worked

The refinement cycle succeeded because disagreements were treated as empirical observations rather than coder errors. Each disagreement was localized, converted into a candidate grammar rule, prospectively tested, and either retained or discarded. This transforms protocol development from expert opinion into an iterative measurement process, the property that most distinguishes this work from a conventional reliability study.

### 4.2 Why graph-level reproducibility matters

A reasonable objection is that if the goal is a reproducible score, one could simply compare CGCI values between coders and report their agreement. Score agreement, however, is a summary that cannot say why two coders differ. Because the specification preserves the intermediate graph, disagreement can be localized to a specific representational choice (a cascade that should have been a parallel assessment, a composite variable that was over-decomposed, a clinical step miscoded as a modifier), and each localized disagreement can be converted into an explicit rule and tested. Graph-level reproducibility is thus not merely a stricter endpoint than score agreement; it is what makes protocol improvement (the refinement cycle) possible. A score comparison alone would tell us that agreement was imperfect without telling us what to change. The stakes are not only methodological: clinical decision-support tools and digital care protocols are built on a translation of the guideline figure into executable logic, and if that translation is not reproducible the resulting software encodes silent, systemic variance in care [1,24]. A reproducible graph representation therefore has implications for software quality assurance and for the consistency of computable guideline implementations, with potential downstream implications for patient safety, beyond its role as a research metric.

### 4.3 Generalization beyond clinical guidelines

Although demonstrated here on guideline algorithms, error-driven grammar induction is not specific to medicine. The same cycle (code independently, localize disagreement, induce a rule, predict where it should help, and test) applies wherever graphs are extracted from structured human-readable artifacts, such as care pathways, business-process diagrams, or causal and influence diagrams [25]. We frame this generalization as potential rather than demonstrated: the present evidence is confined to clinical guidelines, and extension to other domains is an empirical question for future work.

### 4.4 Stopping and the frozen protocol

Iterative refinement requires a principled endpoint, or it continues indefinitely. We adopted the rule that revision continues until successive coding cohorts no longer surface a new disagreement class requiring a new grammar rule, at which point the protocol is frozen. In this study the disagreements observed over successive versions moved from the structural core to the breadth layers and finally to narrow, nameable residuals, with no new structural class emerging, the pattern that motivated freezing the protocol. Declaring an explicit stopping condition, and reporting the residual that remains at freeze, is a stronger scientific position than asserting that a protocol is complete. The Graph Representation Specification is intended to evolve as a versioned technical specification, maintained independently of the CGCI metric [26–32]. The specification is also invariant to guideline content: when a guideline is revised (for example, a future ADA update), the new figures are re-coded under the frozen specification without altering the specification itself.

### 4.5 Relationship to the cognitive-load study

This paper is the measurement foundation for a companion study that interprets CGCI in terms of cognitive load. The interpretation in that study depends on the score being reproducible across coders, which the present work establishes. The two papers score at different granularities — the present paper scores individual decision algorithms, whereas the companion aggregates component counts across an entire guideline — so their absolute CGCI magnitudes are not directly comparable; what transfers between them is the reproducibility of the scoring procedure, not identity of the numbers. Notably, the dominant structural component of the index (the branching-depth product B × D) showed the highest reproducibility, so the between-guideline ordering on which any complexity comparison rests is on the firmest footing. We state the boundary explicitly: these results concern reliability, not construct validity. Whether a higher CGCI corresponds to greater cognitive load in practice is tested in the companion paper; the two results are complementary, not redundant [3,33].

### 4.6 Limitations

The present study demonstrates the feasibility of systematic protocol refinement rather than providing a definitive estimate of inter-rater reliability, and the reproducibility estimates are preliminary. They rest on two coders and four guideline algorithms, so any single pooled reliability coefficient would carry a wide confidence interval; for this reason we report per-guideline agreement and component-level differences rather than a single number. One development cohort revised previously entered codings rather than coding entirely afresh, introducing mild anchoring; breadth agreement, though much improved by explicit rules, remained lower than topological agreement on two guidelines. These limitations motivate a pre-registered confirmatory study that applies the frozen protocol to an expanded set of guidelines with fresh, fully independent coding, and that reports the topology-agreement vector together with limits of agreement rather than a bare coefficient [34,35]. During instrument development the sampling unit is the class of disagreement rather than the individual rater, so expanding the guideline corpus provides more information for protocol refinement than adding coder pairs; that study therefore expands the sampling of guidelines rather than of coders.

## 5. Conclusion

We developed and prospectively evaluated a reproducible method for developing a graph representation specification: disagreement between independent coders is localized, converted into an explicit grammar rule, and prospectively tested for its effect on agreement, with the protocol frozen once no new disagreement class emerges. The resulting specification provides a reproducible foundation for the Clinical Guideline Complexity Index and, more broadly, illustrates how disagreement-driven refinement can mature graph-based measurement systems beyond expert consensus alone.

## Data Availability

All data produced in the present study are available upon reasonable request to the authors

## Declarations

### CRediT authorship contribution statement

Richard V. Milani: Conceptualization, Methodology, Investigation, Writing – original draft, Writing – review & editing, Supervision. Robert M. Bober: Conceptualization, Methodology, Investigation, Writing – review & editing.

### Funding

This research received no specific grant from any funding agency in the public, commercial, or not-for-profit sectors.

### Competing interests

The authors declare that they have no competing interests.

### Ethics

This study did not involve human participants, identifiable human data, or animals; it analyzed published clinical-guideline figures. Institutional review board approval was therefore not required.

### Data and code availability

The scoring engine, the Decision Topology Agreement tool, and the frozen specification (Appendix A) are available from the corresponding author on reasonable request.

## Appendix A. The Graph Representation Specification (frozen protocol v2.6.1)

*This appendix is the NORMATIVE specification of the frozen Graph Representation Specification (v2.6.1). In the event of any discrepancy between the manuscript narrative and implementation details, this appendix is authoritative. No component changes after v2.6.1; earlier versions appear only in the version history (A.9)*.

### A.1 Design principles

Five principles govern the specification and resolve most coding decisions before the detailed rules are consulted:

- Prefer one canonical representation per visual motif.
- Preserve the figure’s explicit structure; encode what is drawn.
- Do not infer clinical knowledge absent from the figure.
- Separate topology from breadth (structure from inputs).
- Resolve disagreement through empirical refinement rather than expert opinion.

### A.2 Scoring model

The index is computed from a valid graph as **CGCI = (B × D) + V + M**. The number of terminal pathways (N_terminal) and the topological-phase indicator (T) are recorded and reported alongside the composite, not summed into it.

| Term | Definition |
| --- | --- |
| **B: branch points** | Count of decision nodes with $\geq 2$ distinct successors. |
| **D: decision depth** | Two-level equal-weight mean of the number of decision nodes on root-to-leaf paths, computed after removing loop_back edges. |
| **V: variables** | Deduplicated union of variables across all decision nodes. |
| **M: modifiers** | Modifiers deduplicated within each figure, then summed across figures. |
| **N_terminal** | Deduplicated union of terminal_class values (reported, not summed). |
| **T: topological phase** | 3 if a cycle (loop_back) or multiple intake nodes; 2 if reconvergence or a cross-figure edge; otherwise 1. |

### A.3 Ontology

A graph is one or more algorithms, each a set of nodes and directed edges. Node types are **decision**, **action**, and **terminal**. Variables attach to decisions only. Modifiers attach to decisions, actions, or terminals (a terminal that states a qualified recommendation carries its qualifiers as modifiers). A terminal carries a terminal_class and has no outgoing edges. An edge is a source-to-destination pair with an optional label and an optional loop_back flag. Unlike variables, modifiers qualify recommendations regardless of node type; a terminal that states a qualified recommendation may therefore legitimately carry modifiers. A motif is a recurring local graph topology with one canonical encoding (A.4).

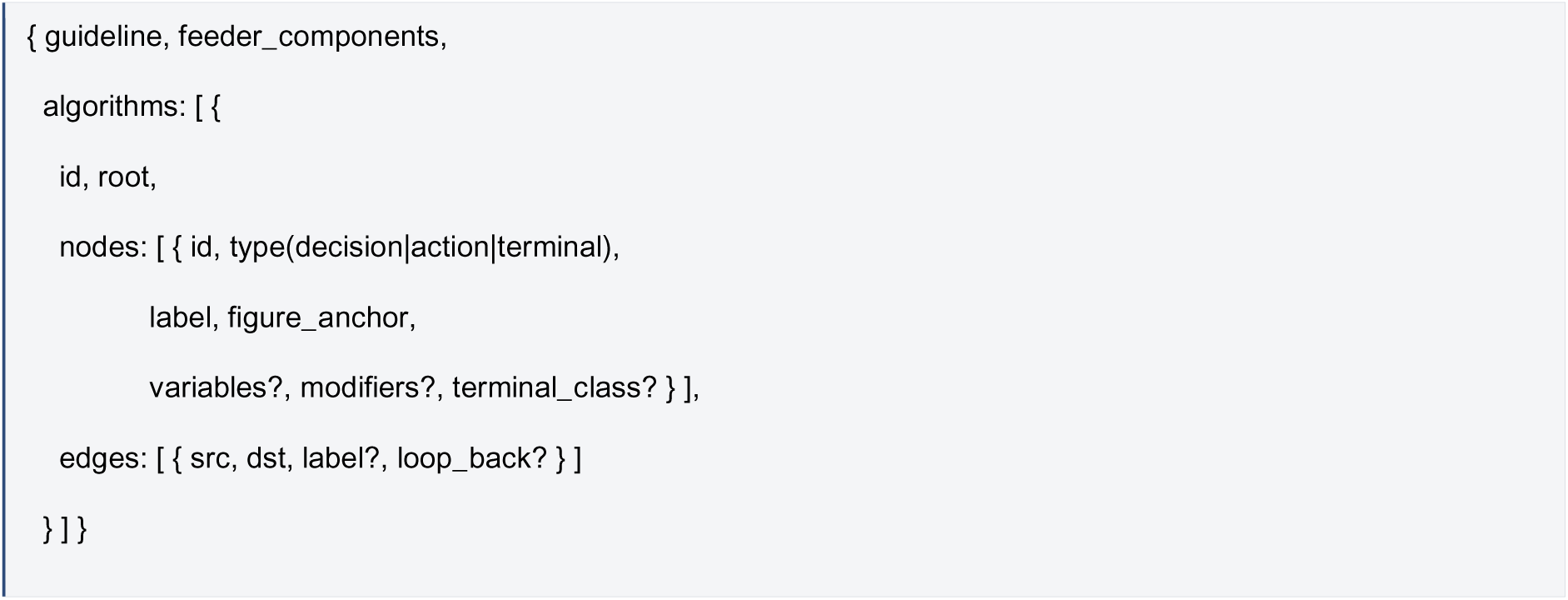

### A.4 Motif catalogue

Coders classify the motif of a figure region first, then encode to its canonical form. Five motifs are defined.

| Motif | Recognition test | Canonical encoding | Counterexample<br>(NOT this motif) |
| --- | --- | --- | --- |
| **Sequential gate** | Indicators tested in order; a 'no' routes to the next distinct step (they do not co-occur). | Chain of binary decisions; C3 applies (add the implicit NO edge). | Parallel assessment: indicators that co-occur in one patient. |
| **Parallel assessment (D6)** | Can multiple indicators be simultaneously true for one patient without excluding one another, AND does the figure evaluate each independently? | ONE assessment decision carrying each indicator as a variable, branching in parallel. C3 does NOT apply. | Sequential gate: mutually exclusive, order-dependent indicators. |
| **Shared rail (C7)** | Several outcomes fed from one shared connector with inward-only arrowheads. | Parallel terminals from the common parent; no box-to-box edges. | A funnel: boxes chained into one another with box-to-box edges. |
| **Threshold ladder (C5)** | Mutually exclusive bands of a single continuous input routing to the same TYPE of step. | ONE categorical decision (unless arrows show sequential testing). | Repeated thresholds: the same cutoff re-tested at separate boxes (D4). |
| **Reassessment loop** | An arrow returns to an earlier box after a time interval or reassessment. | A loop_back edge (sets T = 3); removed before depth is computed. | Forward-only progression, no return arrow. |

#### Distinguishing test (parallel vs. sequential)

Can multiple indicators be simultaneously true for one patient without excluding one another, and does the figure evaluate each independently? YES → parallel assessment (one node, D6). NO → sequential gate (separate decisions, C3 applies).

### A.5 Conventions

Conventions disambiguate coding between the grammar and the validator. The load-bearing conventions:

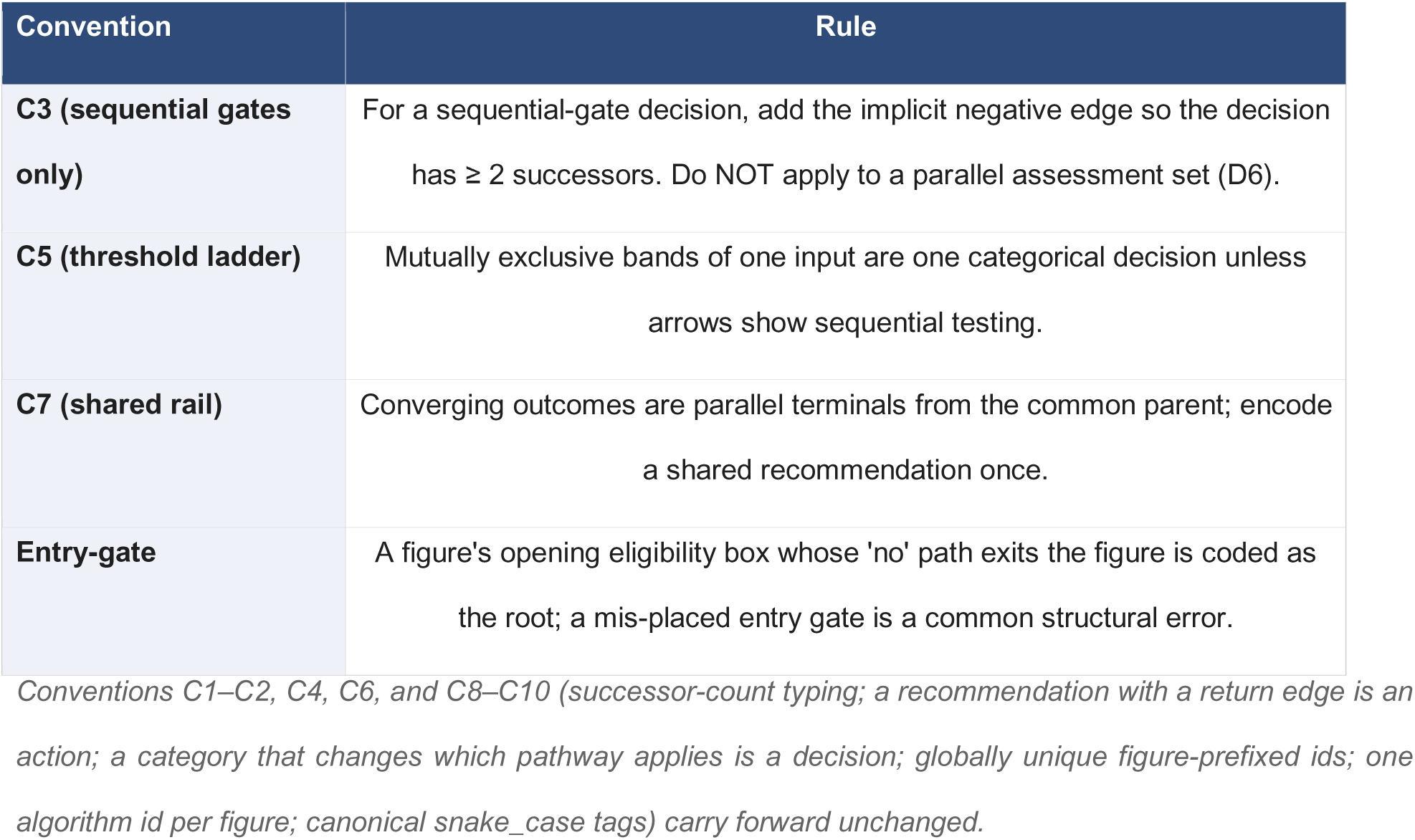

| Convention | Rule |
| --- | --- |
| **C3 (sequential gates only)** | For a sequential-gate decision, add the implicit negative edge so the decision has $\geq 2$ successors. Do NOT apply to a parallel assessment set (D6). |
| **C5 (threshold ladder)** | Mutually exclusive bands of one input are one categorical decision unless arrows show sequential testing. |
| **C7 (shared rail)** | Converging outcomes are parallel terminals from the common parent; encode a shared recommendation once. |
| **Entry-gate** | A figure's opening eligibility box whose 'no' path exits the figure is coded as the root; a mis-placed entry gate is a common structural error. |

*Conventions C1–C2, C4, C6, and C8–C10 (successor-count typing; a recommendation with a return edge is an action; a category that changes which pathway applies is a decision; globally unique figure-prefixed ids; one algorithm id per figure; canonical snake\_case tags) carry forward unchanged.*

### A.6 Decomposition rules

| Rule | Specification |
| --- | --- |
| **D1** | One decision node per point where the figure splits patients into $\geq 2$ pathways. |
| **D2** | Mutually exclusive, sequentially-tested criteria that route to different recommendations are separate decisions (sequential-gate motif only). Co-occurring indicators use D6. |
| **D3** | Mutually exclusive bands routing to the same TYPE of step are one categorical decision. |
| **D4** | Do not duplicate a re-tested threshold; a threshold is a new decision only if drawn as a separate box leading to a different action. |
| **D5: Semantic Decomposition** | Count one variable per input the figure labels. Do not expand a single composite term (e.g., 'health status') or a validated score (e.g., PREVENT, CHA2DS2-VASc) into constituents. Eligibility thresholds are variables of the candidacy decision. |
| **D5b: Modifier Counting** | A modifier qualifies a recommendation (dose, intensity/efficacy tier, numeric target, contraindication, monitoring cadence, population qualifier); count one per printed tier, not per drug. A separate clinical step (risk calculation, screening, testing, referral, procedure) is an action, not a modifier. Modifiers may sit on decisions, actions, or terminals; variables only on decisions. |
| **D6: Assessment Topology** | A set of indicators that can co-occur and are each evaluated independently is one parallel-assessment decision carrying each indicator as a variable; C3 does not apply. |
| **Rule D6 is the defining rule of the specification.**<br>It emerged directly from the calibration cycle: the disagreement class that D6 resolves (co-occurring indicator panels) is what the v2.4 inflation localized, and D6 is the rule whose prospective prediction was confirmed (Results, Section 3.1). |  |

### A.7 Validator (hard rules)

A graph is rejected unless all hold: node ids are globally unique; every edge references an existing node; every node is reachable from the root; terminals have no outgoing edges and carry a terminal_class; variables appear only on decisions; every decision has ≥ 2 distinct successors (after C3); and a cycle appears only as a loop_back edge. Soft flags and informational outputs are implementation-dependent and are therefore omitted from the frozen protocol; only the hard rules above are normative.

### A.8 Reproducibility endpoint (Decision Topology Agreement)

Agreement is computed by contracting each graph to its decision nodes and comparing three components: decision-count agreement, decision-node match rate, and decision-order agreement. Decision nodes are matched by an anchor-first, otherwise label-similarity, optimal (minimum-cost) assignment; the procedure is deterministic and invariant to coder order. Frozen parameters: label similarity = 0.60 × token Jaccard + 0.40 × sequence ratio on canonicalised labels; acceptance threshold τ = 0.40; figure-block normalisation so punctuation differences in ids do not block a match; and an exact figure_anchor match overrides label similarity. Breadth agreement (variables and modifiers) is reported separately.

### A.9 Version history

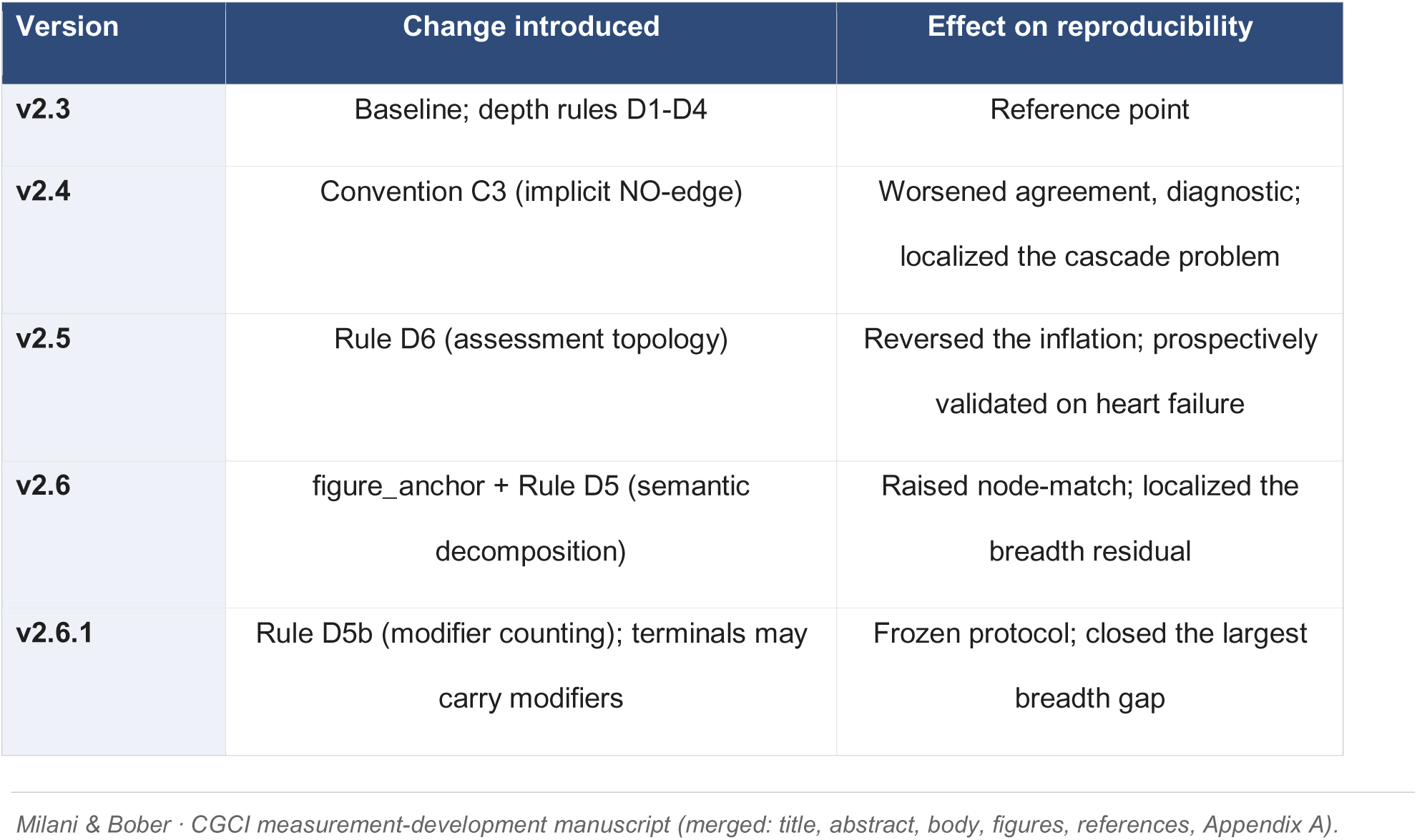

| Version | Change introduced | Effect on reproducibility |
| --- | --- | --- |
| **v2.3** | Baseline; depth rules D1-D4 | Reference point |
| **v2.4** | Convention C3 (implicit NO-edge) | Worsened agreement, diagnostic; localized the cascade problem |
| **v2.5** | Rule D6 (assessment topology) | Reversed the inflation; prospectively validated on heart failure |
| **v2.6** | figure_anchor + Rule D5 (semantic decomposition) | Raised node-match; localized the breadth residual |
| **v2.6.1** | Rule D5b (modifier counting); terminals may carry modifiers | Frozen protocol; closed the largest breadth gap |

## Notes

### Competing Interest Statement

The authors have declared no competing interest.

